# Time to diagnosis and long-term outcomes for adults presenting with breathlessness

**DOI:** 10.1101/2024.02.19.24302618

**Authors:** Urvee Karsanji, Claire A Lawson, Emily Petherick, Kamlesh Khunti, Gillian E. Doe, Jennifer K Quint, Alex Bottle, Michael C Steiner, Rachael A Evans

## Abstract

**Background:** There are known delays to diagnosis for diseases which commonly present with chronic breathlessness, but the subsequent impact is unknown. For adults presenting with breathlessness, we investigated the time taken to achieve an explanatory diagnosis, and associations with unplanned hospitalisation and mortality.

**Methods:** A retrospective cohort study using the UK CPRD was conducted involving adults with a first-recorded code for breathlessness and no pre-existing cardiorespiratory disease. We documented whether an explanatory diagnosis was recorded after the first code of breathlessness within two years and during all follow-up, and the time to diagnosis. Cox regression (adjusted) was used to investigate the associations with unplanned hospitalisation and mortality.

**Results:** 101369 adults were included with a first-recorded code for breathlessness. After two-years, 43394 (43%) adults received a recorded explanatory diagnosis and had a higher risk of unplanned hospitalisation (1.25 [1.19-1.31]) and mortality (2.06 [1.60-2.65]) compared to adults without a diagnosis. Overall, 66909 (66%) adults received a recorded diagnosis during a median of 5-years follow-up. Adults that received a recorded diagnosis after ≥6 months had worse outcomes of unplanned hospitalisation (6-24 months: 1.01 [0.94-1.08]; ≥24 months: 1.13 [1.06-1.20]) and mortality (6-24 months: 3.38 [2.21-5.18]; ≥24 months: 10.80 [7.46-15.70]).

**Conclusion:** We describe a sub-group of adults coded for breathlessness but without an explanatory diagnosis with better outcomes. However, in adults with an explanatory diagnosis waiting beyond six months was associated with worse outcomes. Diagnostic pathways for chronic breathlessness need to differentiate between these two groups and achieve earlier diagnosis in those at higher risk.

**Key messages:** *What is already known on this topic?:*

< Delays to diagnosis exist for chronic cardiorespiratory diseases, but the impact of these delays on future hospitalisation and mortality risk are unknown.

*What this study adds:*

< Over a median follow-up of 5 years, 1 in 2 people with breathlessness had an unplanned hospital admission and 11% died.
< We identify a group of patients with a breathlessness code who did not receive a diagnosis but overall had better outcomes than those with an explanatory diagnosis.
< We also report novel findings that for adults who receive an explanatory diagnosis for breathlessness, waiting beyond six months to receive a diagnosis is associated with an increased risk of future unplanned hospital admission and all-cause mortality.

*How this study might affect research, practice or policy:*

< Further research is needed to prioritise investigations early for patients presenting with chronic breathlessness with increased risk of underlying cardiorespiratory disease.
< Diagnostic breathlessness pathways may improve the time to diagnosis and therefore improve longer term outcomes.
< Where an underlying causative diagnosis of cardiorespiratory disease is not identified, outcomes appear better, and attention can be focused on reassurance and symptom management.

## Introduction

Breathlessness is a common presenting symptom for long-term conditions such as chronic heart and lung diseases (1) and increases with age (2). Although chronic breathlessness should raise concerns about underlying cardiorespiratory disease, there are a wide range of other possible diagnoses and contributing factors including breathing pattern disorder, anaemia, anxiety and obesity (3).

The number of people with chronic diseases such as heart failure and chronic obstructive pulmonary disease (COPD) is set to rise due to an ageing population. Both these cardiorespiratory diseases carry a high risk of morbidity and mortality and can often go undiagnosed until patients have an emergency admission. Delays in diagnosis for COPD have been shown to be multifactorial with factors relating to patient-related symptoms, healthcare provider factors and heterogeneity in the disease itself (4).

Missed opportunities for COPD diagnosis in UK primary and secondary care revealed 85% patients in the cohort having a delay in diagnosis of up to 5 years (5). Similarly, a recent online survey consisting of 600 patients reported similar findings for interstitial lung disease (ILD), with 43% of patients reporting delays of ≥1 year and 19% of patients reporting delays of ≥3 years (6). These diagnostic delays occur despite evidence-based guidelines, due to being disease-specific rather than symptom based i.e. how the person seeks help from healthcare. The National Institute for Health and Care Excellent (NICE) (7) and NHS England (8) recommend investigations to be performed to determine the cause of breathlessness but without a specific pathway or time-frame.

Whilst delays in diagnosis are known, the longer-term impact on future mortality and healthcare utilisation remains unknown. We therefore aimed to document explanatory diagnoses recorded in the primary care record associated with chronic breathlessness and to investigate the relationship between time to diagnosis from first presentation and subsequent mortality and hospitalisation.

## Methodology

### Study design and population

A retrospective cohort study was conducted using routinely collected healthcare data from the UK Clinical Practice Research Datalink (CPRD-GOLD) database between 1 January 2007 and 31 December 2017. 36 Read codes were obtained from Keele University Research Institute for Primary Care and Health Sciences and from a publication by Watson *et al*. (9) (see **Appendix**) to identify breathlessness. All adults with a first-recorded code for breathlessness during the study period eligible for linkage to Hospital Episode Statistics (HES) Admitted Patient Care (APC), Office for National Statistics (ONS) and the 2015 English Index of Multiple Deprivation (IMD) databases were included. The study cohort consisted of all eligible adults with a first-recorded code for breathlessness recorded in their primary care records during the study period.

#### Inclusion criteria

< Patients of acceptable data quality as defined from CPRD and those that were alive and registered between 1 January 2007 and 31 December 2017 (10).
< Patients aged ≥18 years at the start of the study index period (1 January 2007).
< Patients with a first-recorded code for breathlessness during the study index period.
< Patients with at least 12-months data prior to entry into the study cohort and with at least 12-months follow-up data after first-recorded code for breathlessness.
< Patients eligible for linkage to HES APC, ONS and IMD databases.

#### Exclusion criteria

< Patients with a pre-existing code for COPD, heart failure, interstitial lung disease (ILD) or asthma before the study index period in CPRD and HES databases (codes for any asthma event occurring more than 10 years prior to the index date were ignored).

There is no specific code for chronic or persistent breathlessness, so a surrogate strategy was used to exclude patients presenting with acute breathlessness relating to an acute condition. Adults with an acute respiratory infection (upper respiratory tract infection (URTI), lower respiratory tract infection (LRTI) or pneumonia) coded on the same day as breathlessness were considered acute (Read codes can be found in the **Appendix**).

### Outcomes

Diagnoses were defined using Read codes (11) in primary care and International Classification of Disease 10^th^ revision (ICD-10) codes (12) in secondary care. Relevant diagnoses were split into three categories: cardiac conditions (467 codes), pulmonary conditions (253 codes), and other non-cardiorespiratory conditions (643 codes). A full list of codelist sources, Read and ICD-10 codes for diagnoses can be found in the **Appendix**.

Longer-term outcomes of unplanned hospital admission were obtained through HES data and all-cause mortality through ONS data. Only unplanned (non-elective) admissions were considered as the outcome of interest, all elective (planned) admissions were not accepted. Date of admission was used as the first unplanned hospital admission following first-recorded code for breathlessness. For adults that received a first-recorded code for breathlessness on the same day as an admission to hospital, the subsequent unplanned hospital admission was taken.

### Statistical analysis

For baseline characteristics, demographics of the population were assessed. The burden of comorbidities at baseline was obtained by applying the Cambridge Multimorbidity Score (13). The study focused on two components of analysis (see **Figure 1**):

**Figure 1.**
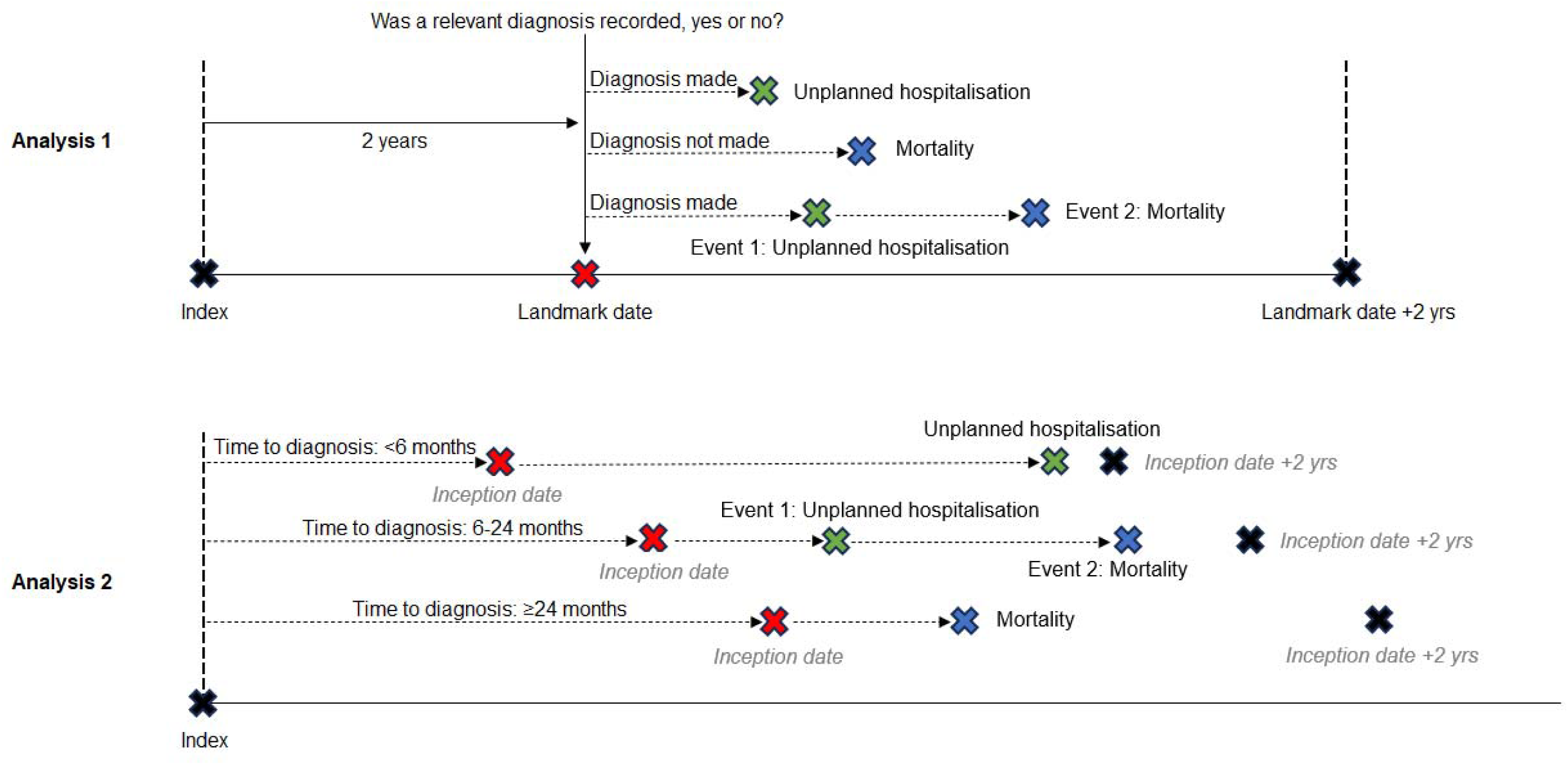
Study period diagram showing the components of Analysis 1 and Analysis 2 with

1. <u>Effect of receipt of an explanatory diagnosis or not</u> A 2-year landmark date was set after first-recorded code for breathlessness (index) to assess whether an explanatory diagnosis associated with breathlessness had been recorded or not. Comparison of adults with and without a recorded diagnosis at landmark date on subsequent outcomes of unplanned hospital admission and all-cause mortality were assessed. Inception date for this analysis was set as the 2-year landmark date after presentation with breathlessness. Outcomes were assessed in the following two years from landmark date.
2. <u>Effect of time to diagnosis from first code of breathlessness</u> During a median of 5 years follow up, all adults that received a recorded explanatory diagnosis following first-recorded code for breathlessness were described and categorised into ‘time to diagnosis’ categories: ‘<6 months’, ‘6-24 months’ and ‘≥24 months’. The association between time to reaching that diagnosis and subsequent outcomes of unplanned hospital admissions and all-cause mortality was investigated. Inception date for this analysis was the date of recorded diagnosis.

In the above two components, Cox regression was used to investigate the association between 1) receiving a recorded diagnosis or not and outcomes of unplanned hospital admission and all-cause mortality and 2) time to diagnosis and outcomes of unplanned hospital admission and all-cause mortality. Models were adjusted for the following covariates: sex, age, neighbourhood deprivation, BMI, smoking status, ethnicity, the number of comorbidities and prior hospital admission. All these covariates were part of an unadjusted analysis and were selected a priori, and hence were entered into a multivariable model together. A prior admission was defined as an unplanned hospital admission that occurred between the first recorded code for breathlessness and landmark date. The reference group for comparisons was set as adults that did not receive a recorded diagnosis in Analysis 1 and adults that received a recorded diagnosis in <6 months for Analysis 2. For modelling purposes, missing data for covariates of BMI and smoking status were imputed using multiple imputation across 5-folds (14). For adults that received a relevant recorded diagnosis on the same date as first unplanned hospital admission, the next unplanned hospital admission was used as the outcome.

## Results

### Patient characteristics

Health records from 103917 adults presenting with a first-recorded code for breathlessness and no known cardiorespiratory disease were included from CPRD-GOLD during the study period (**Figure 2**). ‘Shortness of breath’ (medcode 4822; Read code 1739) was the most frequent code to capture breathlessness (n = 29204, 29%), followed by the ‘MRC Dyspnoea Scale Grades 1-5’ (n = 16948, 17%). 2495 adults with an acute respiratory infection code on the same day as being coded for breathlessness (URTI = 887; LRTI = 1469; pneumonia = 139) were removed (see **Appendix**). A further 53 patients were excluded due to data anomalies (i.e. having a death date before a recorded diagnosis or unplanned hospital admission) leaving the final cohort of patients to be n = 101369.

**Figure 2.**
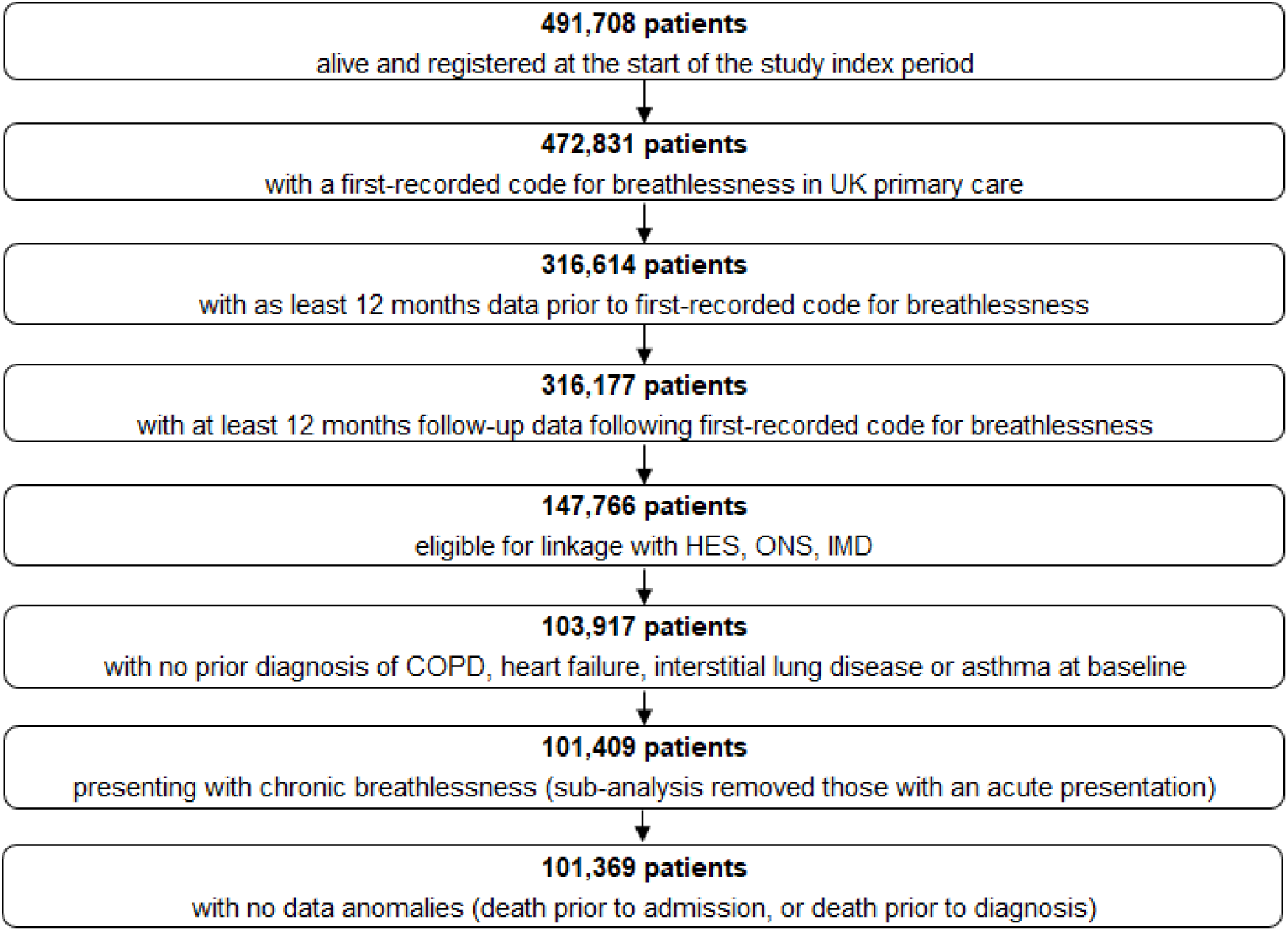
Patient selection flowchart.

The median [IQR] follow-up time from index date (first code of breathlessness) was 5 [3 – 7] years. Patient characteristics at the three different time points are shown in **Table 1**: first-recorded code for breathlessness, landmark date (Analysis 1) and date of diagnosis (Analysis 2). At the time of first-recorded code for breathlessness, over half of the cohort presented with two or more comorbidities at baseline (56%), and just under a fifth presenting with none (19%). The two most common comorbidities amongst the cohort were hypertension (31%) and depression (30%). The majority of missingness at baseline was for BMI (15%) and smoking status (14%).

**Table 1.** Patient characteristics at index (first-recorded code for breathlessness), landmark date and date of diagnosis.

|  | Index | Landmark date |  | Date of diagnosis |  |  |
| --- | --- | --- | --- | --- | --- | --- |
|  | All | No diagnosis | Diagnosis | <6 months | 6-24 months | ≥24 months |
| Number of patients | 101,369 (100.0) | 57,975 (57.2) | 43,394 (42.8) | 28,580 (42.7) | 14,787 (22.1) | 23,542 (35.2) |
| Gender, n (%) |  |  |  |  |  |  |
| Male | 45,312 (44.7) | 25,349 (43.7) | 19,963 (46.0) | 13,776 (48.2) | 6,175 (41.8) | 9,972 (42.4) |
| Female | 56,057 (55.3) | 32,626 (56.3) | 23,431 (54.0) | 14,804 (51.8) | 8,612 (58.2) | 13,570 (57.6) |
| Mean ± SD, Age (years) | 58.1 ± 16.4 | 58.0 ± 16.4 | 62.8 ± 15.9 | 61.6 ± 15.4 | 60.8 ± 16.7 | 65.0 ± 16.5 |
| Ethnicity, n (%) |  |  |  |  |  |  |
| Black | 1,151 (1.1) | 746 (1.3) | 405 (0.9) | 238 (0.8) | 166 (1.1) | 249 (1.1) |
| Mixed/Other | 1,503 (1.5) | 989 (1.7) | 514 (1.2) | 319 (1.1) | 195 (1.3) | 297 (1.3) |
| South Asian | 2,293 (2.3) | 1,467 (2.5) | 826 (1.9) | 495 (1.7) | 331 (2.2) | 459 (2.0) |
| White | 87,596 (86.4) | 48,451 (83.6) | 39,145 (90.2) | 25,797 (90.3) | 13,324 (90.2) | 21,335 (90.5) |
| Unknown/Missing | 8,826 (8.7) | 6,322 (10.9) | 2,504 (5.8) | 1,731 (6.1) | 771 (5.2) | 1,202 (5.1) |
| Mean ± SD, BMI (kg/mass) | 28.5 ± 6.5 | 28.0 ± 5.6 | 28.8 ± 6.4 | 28.6 ± 7.1 | 29.2 ± 7.4 | 28.9 ± 7.0 |
| Not obese (<30) | 63,157 (62.3) | 37,161 (64.1#) | 26,040 (60.0) | 17,742 (62.1) | 8,556 (57.9) | 13,156 (55.9) |
| Obese (≥30) | 22,743 (22.4) | 16,434 (28.4) | 15,113 (34.8) | 9,771 (34.2) | 5,362 (36.3) | 8,787 (37.3) |
| Missing | 15,469 (15.3) | 3,515 (6.1) | 1,666 (3.8) | 1,067 (3.7) | 869 (5.9) | 1,599 (6.8) |
| Smoking Status, n (%) |  |  |  |  |  |  |
| Current smoker | 20,693 (20.4) | 8,884 (15.3) | 17,945 (41.4) | 6,863 (24.0) | 2,558 (17.3) | 3,577 (15.2) |
| Non-smoker | 34,181 (33.7) | 18,402 (52.5) | 10,461 (24.1) | 11,126 (38.9) | 6,887 (46.6) | 11,551 (49.1) |
| Ex-smoker | 41,282 (40.7) | 30,454 (31.7) | 14,498 (33.4) | 10,515 (36.8) | 5,320 (36.0) | 8,381 (35.6) |
| Data Not Entered | 5,213 (5.1) | 235 (0.4) | 490 (1.1) | 76 (0.3) | 22 (0.2) | 33 (0.1) |
| Deprivation, n (%) |  |  |  |  |  |  |
| 1 (least) | 22,968 (22.7) | 13,854 (23.9) | 9,114 (21.0) | 5,981 (20.9) | 3,127 (21.3) | 5,228 (21.2) |
| 2 | 22,229 (21.9) | 12,952 (22.3) | 9,277 (21.4) | 6,111 (21.4) | 3,164 (2.4) | 5,224 (22.2) |
| 3 | 21,128 (20.8) | 12,160 (21.0) | 8,968 (20.7) | 5,905 (20.7) | 3,055 (20.7) | 4,927 (20.9) |
| 4 | 18,280 (18.0) | 10,092 (17.4) | 8,188 (18.9) | 5,351 (18.7) | 2,830 (19.1) | 4,210 (17.9) |
| 5 (most) | 16,764 (16.5) | 8,917 (15.4) | 7,847 (18.0) | 5,232 (18.3) | 2,611 (17.7) | 3,953 (16.8) |
| Comorbidities, n (%) |  |  |  |  |  |  |
| 0 | 19,606 (19.3) | 16,997 (29.3) | 8,595 (19.8) | 4,550 (15.9) | 1,619 (11.0) | 2,482 (10.5) |
| 1 | 24,600 (24.3) | 17,679 (30.5) | 11,519 (26.5) | 6,356 (22.2) | 2,840 (19.2) | 4,679 (19.9) |
| ≥2 | 57,163 (56.4) | 23,299 (40.2) | 23,280 (53.7) | 17,674 (61.8) | 10,328 (69.8) | 16,381 (69.6) |

43394 (43%) adults had received a relevant recorded diagnosis within 2 years of first-recorded code for breathlessness (landmark date). Adults that received a relevant recorded diagnosis by landmark date were on average five years older and had a greater BMI than adults that did not receive a diagnosis. Of the ethnicity data available, 94% of adults that did not receive a diagnosis were White, 1% Black, 2% Mixed/Other and 3% South Asian, and 96% of adults that did receive a diagnosis were White, 1% were Black, 1% were Mixed/Other, and 2% were of South Asian ethnicity. Of the smoking data available, 15% were current smokers, 32% non-smokers and over half of adults with no diagnosis were ex-smokers; whereas there were a greater number of current smokers (19%) and non-smokers (42%) among those who received a diagnosis and fewer ex-smokers (39%). Over half of adults that received a diagnosis had ≥2 existing comorbidities, compared with two-fifths of adults that did not receive a diagnosis. 136 patients died between index and landmark date and were removed from mortality models, and 24044 (23.7%) patients had a prior admission between index and landmark date.

Overall, 66909 (66%) adults received a diagnosis during a median follow-up of 5 years. 41700 (62%) received their primary diagnosis in the community and 25209 (38%) received their primary diagnosis in hospital. Of adults that had a diagnosis made during the median 5-year follow-up, 51859 (77.5%) had a cardiorespiratory diagnosis made and 15050 (22.5%) had a non-cardiorespiratory diagnosis. A single relevant diagnosis was recorded for 29989 (44.8%) adults and 36920 (55.2%) adults had multiple diagnoses recorded. Asthma and COPD were the two most prevalent diagnoses recorded amongst the cohort (17%) followed by cardiac arrhythmias (16%) and then anxiety (14%). Pre-existing diagnoses (from the relevant conditions investigated) were present for a third of the cohort, with the most prevalent being anxiety (28.2%), depression (16.9%) and coronary heart disease (14.1%).

The median [IQR] time to diagnosis was 316 days [31 – 1157]. 28580 (43%) adults received a relevant diagnosis in <6 months, 14787 (22%) in 6-24 months and 23542 (35%) in ≥24 months. Adults that received a diagnosis in ≥24 months were on average five years older than adults that received a diagnosis in <24 months. Of those with smoking data available, over a third of adults are ex-smokers in each time to diagnosis category, and there were more non-smokers among adults that took ≥6 months to receive a diagnosis (48%) compared with those that took <6 months (39%). 11% adults that received a diagnosis in ≥6 months had no comorbidities at baseline compared with 16% adults diagnosed in <6 months. **Table 2** shows the explanatory diagnoses recorded in the cohort within 6 months, 2 years and a median follow-up of 5 years.

**Table 2.** Relevant diagnoses recorded within 6 months, 2 years and 5 years from first-recorded presentation with breathlessness.

| Diagnoses | 6-months |  | 2-years |  | 5-years* |  |
| --- | --- | --- | --- | --- | --- | --- |
|  | N | % | N | % | N | % |
| Cardiac | 11,992 | 11.83 | 21,159 | 20.87 | 51,018 | 50.33 |
| Heart failure | 2,076 | 2.05 | 3,240 | 3.20 | 7,937 | 7.83 |
| Left ventricular hypertrophy | 687 | 0.68 | 1,565 | 1.54 | 4,686 | 4.62 |
| Right ventricular hypertrophy | 39 | 0.04 | 68 | 0.07 | 102 | 0.10 |
| Coronary heart disease | 2,666 | 2.63 | 5,211 | 5.14 | 11,098 | 10.95 |
| Cardiomyopathy | 377 | 0.37 | 676 | 0.67 | 1,278 | 1.26 |
| Valvular disease | 1,765 | 1.74 | 2,996 | 2.96 | 8,278 | 8.17 |
| Cardiac arrhythmias/AF | 4,124 | 4.07 | 6,922 | 6.83 | 16,390 | 16.17 |
| Pericardial disease | 169 | 0.17 | 300 | 0.30 | 863 | 0.85 |
| Congenital heart disease | 89 | 0.09 | 181 | 0.18 | 386 | 0.38 |
| Pulmonary | 16,423 | 16.20 | 24,213 | 23.89 | 47,798 | 47.15 |
| COPD | 8,007 | 7.90 | 10,311 | 10.17 | 16,885 | 16.66 |
| Asthma | 5,624 | 5.55 | 9,184 | 9.06 | 17,052 | 16.82 |
| ILD | 224 | 0.22 | 418 | 0.41 | 923 | 0.91 |
| Lung cancer | 113 | 0.11 | 210 | 0.21 | 1,069 | 1.05 |
| Pleural effusion | 968 | 0.95 | 1,539 | 1.52 | 5,507 | 5.43 |
| Bronchiectasis | 316 | 0.31 | 744 | 0.73 | 2,080 | 2.05 |
| Thromboembolic disease | 848 | 0.84 | 1,096 | 1.08 | 1,565 | 1.54 |
| Pulmonary hypertension | 209 | 0.21 | 445 | 0.44 | 1,628 | 1.61 |
| Neuromuscular disorders | 20 | 0.02 | 35 | 0.03 | 58 | 0.06 |
| Chest wall deformities | 94 | 0.09 | 231 | 0.23 | 1,031 | 1.02 |
| Other non-cardiorespiratory | 8,779 | 8.66 | 20,154 | 19.88 | 47,860 | 47.21 |
| Anaemia | 2,647 | 2.61 | 5,067 | 5.00 | 12,137 | 11.97 |
| Anxiety | 2,673 | 2.64 | 6,539 | 6.45 | 14,650 | 14.45 |
| Depression | 1,205 | 1.19 | 3,465 | 3.42 | 6,706 | 6.62 |
| Obesity | 1,850 | 1.83 | 4,414 | 4.35 | 12,895 | 12.72 |
| Dysfunctional breathing | 404 | 0.40 | 669 | 0.66 | 1,472 | 1.45 |
| No diagnosis | 72,771 | 71.79 | 57,975 | 57.19 | 34,460 | 33.99 |

\*Some patients had multiple diagnoses made during the median follow-up of 5 years, hence percentages do not sum up to 100%. Of adults that had a diagnosis made during the median 5-year follow-up, 51859 (77.51%) had a cardiorespiratory diagnosis made and 15050 (22.49%) had a non-cardiorespiratory diagnosis.

### The association between receiving a diagnosis or not and longer-term outcomes

The median [IQR] follow-up time from landmark date for an unplanned hospital admission was 841 days [320 – 1815] and 1192 days [467 – 2030] for mortality. Within 2 years of landmark date, there were more unplanned hospital admissions among adults that received an explanatory diagnosis compared with adults that did not (10860 (25%) vs 9187 (16%)) and a greater number of deaths (755 (2%) vs 286 (0.5%)). Adults who received a recorded explanatory diagnosis by landmark date (within two-years from first recorded code of breathlessness) had a greater risk of unplanned hospital admission (1.75 [1.70 – 1.80], *p*<0.001) and all-cause mortality (3.77 [3.29 – 4.31], *p*<0.001) within the subsequent 2 years compared with adults who did not receive a diagnosis (**Figure 3**). Adjusted Cox regression showed similar results, with adults who received a relevant recorded diagnosis having a higher risk of unplanned hospital admission and all-cause mortality within 2 years compared with adults that did not receive a diagnosis (unplanned hospital admission: 1.25 [1.19 – 1.31], *p*<0.001); all-cause mortality 2.06 [1.60 – 2.65], *p*<0.001).

**Figure 3.**
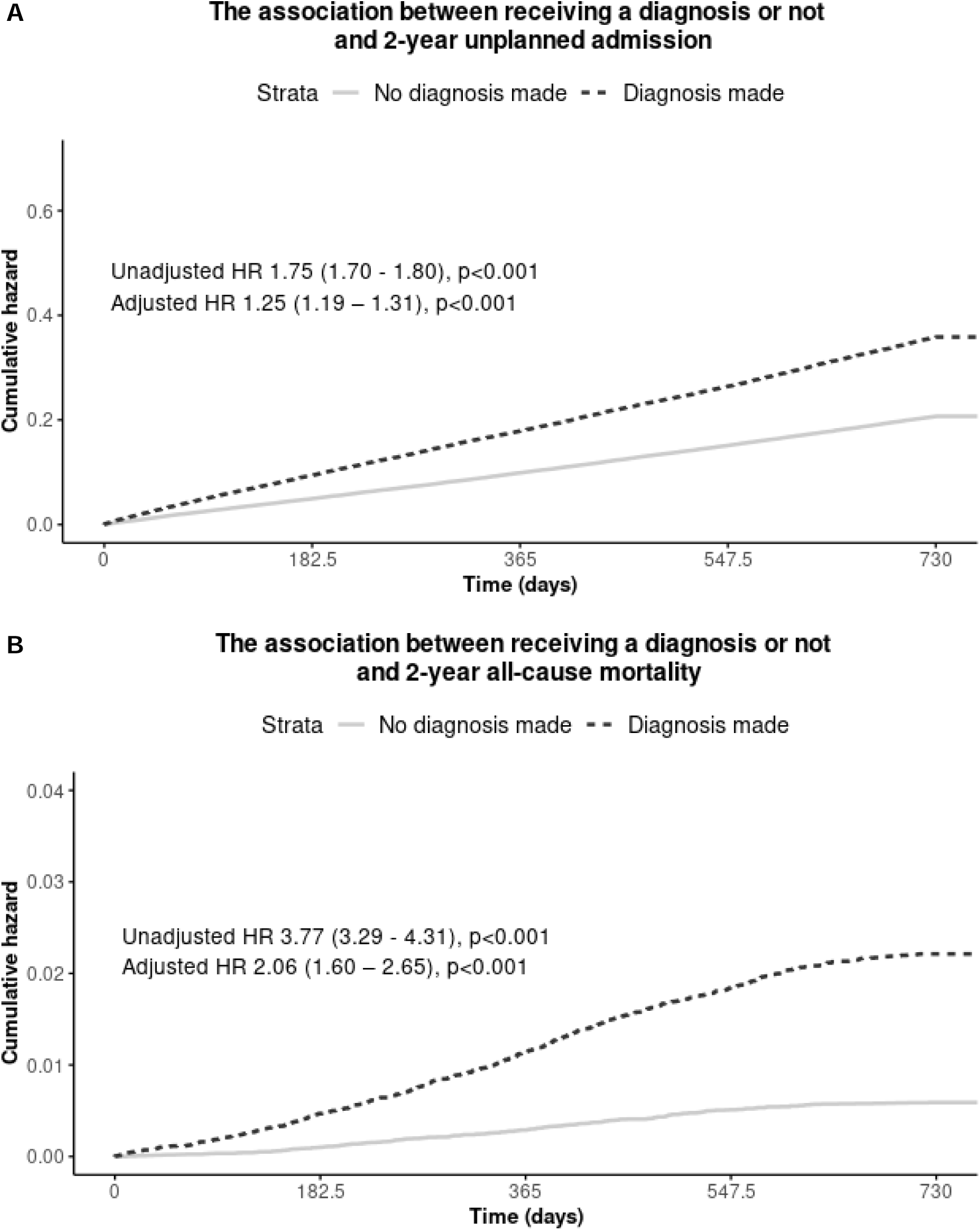
Unadjusted Kaplan-Meier curves showing the effect of receiving a diagnosis or not on outcome. **A** shows an outcome of 2-year unplanned hospital admission. **B** shows an outcome of 2-year all-cause mortality. Solid line = adults that did not receive a relevant diagnosis in 2 years of landmark date (reference); Dotted line = adults that received a relevant diagnosis in 2 years of landmark date.

### The association between time to diagnosis and outcome

The median [IQR] follow-up time from date of diagnosis for an unplanned hospital admission was 827 days [368 – 1674] and 1240 days [518 – 2131] for mortality. There were fewer unplanned hospital admissions and deaths for adults that had a recorded diagnosis in <6 months (40% and 1%, respectively) compared to 6-24 months (43% and 2%, respectively) and ≥24 months (43% and 6%, respectively). Adults that took longer to receive a diagnosis had a higher risk of unplanned hospital admission and all-cause mortality compared with adults who received a diagnosis in <6 months (**Figure 4**). Adjusted Cox regression showed similar risk estimates for unplanned hospital admission (6-24 months: 1.01 [0.94 – 1.08], *p*=0.828; ≥24 months: 1.13 [1.06 – 1.20], *p*<0.001) and all-cause mortality in 2 years (6-24 months: 3.38 [2.21 – 5.18], *p*<0.001; ≥24 months: 10.80 [7.46 – 15.70], *p*<0.001) compared with adults diagnosed in <6 months.

**Figure 4.**
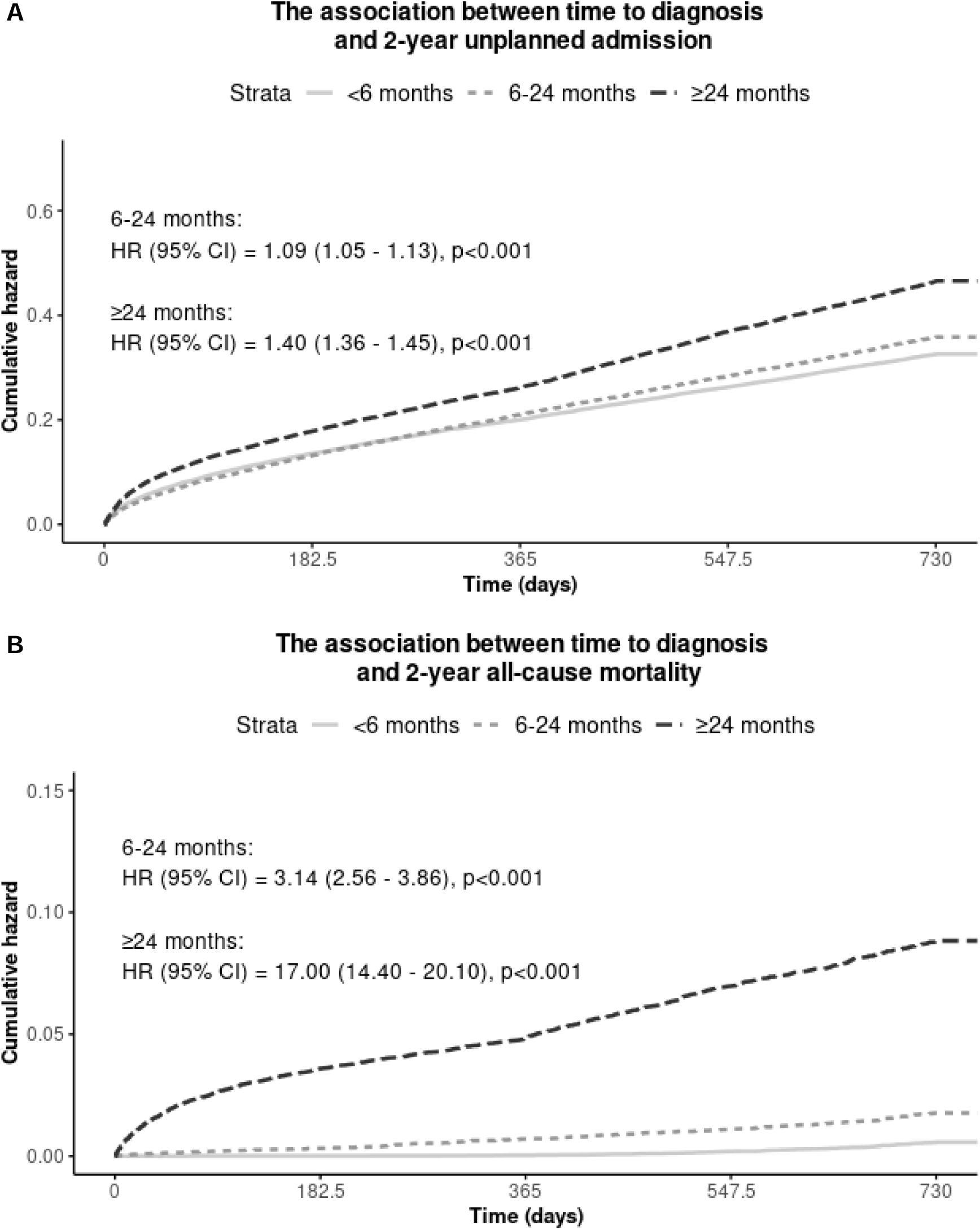
Unadjusted Kaplan-Meier curves showing the association between time to diagnosis and outcome. **A** shows an outcome of 2-year unplanned hospital admission. **B** shows an outcome of 2-year all-cause mortality. Solid line: diagnosis made <6 months; Small dash: diagnosis made within 6-24 months; Large dash: diagnosis made ≥24 months.

## Discussion

We report a large cohort study of 101369 adults with a first recorded code of breathlessness using UK primary care electronic healthcare records and highlight significant healthcare utilisation and mortality associated with this symptom. We report for the first time two distinct groups with differing outcomes; adults with an explanatory diagnosis (predominantly chronic cardiorespiratory disease) associated with worse outcomes and a group or adults who present with breathlessness but without an explanatory diagnosis who overall appear to have a better prognosis. Less than 6 in 10 adults with a code for breathlessness received an explanatory diagnosis within two years of seeking healthcare.

Approximately 4 in 10 adults were diagnosed at an unplanned hospital admission, indicating a more severe condition which is a concern for the individual as well as the healthcare system. Furthermore, we report for the first time to our knowledge, that the time taken from the first presentation with breathlessness to receive an explanatory diagnosis was associated with worse outcomes (higher mortality and increased unscheduled hospital admissions).

Other reports typically start at the time of diagnosis for a particular condition such as COPD, heart failure asthma and have looked back for the first recorded code of breathlessness and shown delays in diagnosis (5, 6). We have expanded this approach by taking the symptom of breathlessness as the central point, in line with how a person seeks healthcare, and worked forwards providing new information on the group without an explanatory diagnosis who appear to do better and new information on the effects of time to diagnosis for the symptom of breathlessness irrespective of underlying diagnosis. In adults who received a relevant recorded diagnosis (predominantly cardiorespiratory diseases) during follow-up, the time to diagnosis was positively associated with unplanned hospital admissions and mortality. We suggest this analysis highlights the importance of timely diagnosis of underlying cardiorespiratory conditions and suggests earlier diagnosis offers meaningful therapeutic opportunity. The deployment of symptom-based diagnostic pathways in primary care aiming to achieve prompt diagnosis and treatment, has the potential therefore to improve outcomes (15, 16).

There are several individual characteristics that were associated with better outcomes despite no diagnosis being made after breathlessness being recorded, including younger adults, lower BMI, non-smokers and fewer comorbidities. This highlights that not all breathlessness codes are associated with poor outcomes, particularly for individuals where no specific diagnosis is achieved. There appears to be a cohort of patients with symptoms of chronic breathlessness that need symptom control similar to chronic cough or pain, who do not necessarily develop severe disease leading to hospitalisation or premature mortality. Having codes/pathways which can differentiate this future risk would help prioritise/stratify healthcare to the greatest need.

Landmark analysis indicated that adults with a recorded diagnosis after presentation with breathlessness have worse all-cause mortality and higher rates of unscheduled hospital admission than those without an explanatory diagnosis. We suggest that the absence of a coded underlying cause (principally chronic cardiorespiratory disease) for some patients indicates a more benign condition (albeit not necessarily regarding distress to the individual) potentially due to causes such as breathing pattern disorder or anxiety. We do not infer however, that the symptom itself is less severe as this cannot easily be determined from the primary care record nor that the symptom itself does not require therapeutic intervention. This supports the need for symptom-based diagnostic pathways for earlier diagnosis and treatments.

### Strengths and limitations

The key strengths of the CPRD relay to breadth of coverage, size, long-term follow-up, representativeness and quality of data (17). This allows associations to be explored and results will hold a higher statistical power due to the large sample size, which holds better with rarer diseases and exposures. Comparing CPRD data with the UK census in 2011 (18) showed valid representativeness of CPRD patients for age, gender, and ethnicity. Through linkage to secondary data via HES, IMD and the ONS, mortality and lifestyle variables are captured, which can give a better insight into patient characteristics. In addition, through the implementation of the Quality Outcomes Framework (19), data quality has been promoted for most chronic conditions (20, 21).

The Read codes used do not distinguish whether patients have an acute or chronic presentation with breathlessness. Based on clinical input, it was decided that if a code for an acute respiratory infection (URTI, LRTI or pneumonia) had been entered on the same date as first-recorded code for breathlessness then this represented an acute presentation. Without confirmation of the free text in the medical record entered by the GP, or a new code being created for ‘acute breathlessness’ and ‘chronic breathlessness’, there is no guarantee that these patients are presenting with chronic breathlessness. There is a need for more well-defined coding terminology for breathlessness to differentiate acute from chronic or persistent, and whether the symptom continues despite optimal medical management.

In recent years, there has been discussion about defining breathlessness as a syndrome (22, 23), recognising that there are a proportion of patients who are on treatment and management plans but have ongoing symptoms of breathlessness who are at greater need of support and who have a worse prognosis. The MRC dyspnoea scale assesses the functional impact of breathlessness and is associated with prognosis in the general population and in long term conditions (24, 25). However, it only tends to be used for chronic respiratory disease. Most codes do not differentiate the severity of the experience of breathlessness and simple descriptors do not overall differentiate between different underlying conditions (26).

Despite identifying a large cohort of adults, we believe this to be an underestimate. It is not routine practice for primary care practitioners to code symptoms. From the 36 Read codes used to identify breathlessness, 18 of these codes were rarely used (<1%). Watson *et al*. (9) devised a codelist for breathlessness using Read codes, which was the basis for obtaining our cohort. These codes have not been validated, but were cross-referenced with the codes used by Chen *et al*. (22). With coding inconsistencies in the UK and no set guidelines on how to code effectively for research, GPs differ in coding styles, and this may have affected the number of patients identified to be presenting with breathlessness during the study index period. At the time of consultation, it is unknown whether only the primary reason for the consultation is recorded in the medical record or whether this is the case for all other signs and symptoms. If GPs struggle to reach a diagnosis, then the symptom could perhaps get overlooked and under-recorded. Jensen *at al.* (23) also found that if symptoms were not accurately reflective of medical codes then free text could be used instead. For research studies, this would mostly be overlooked due to free text not being available unless requested from general practice in primary care; free text in secondary care is not possible so only the terms captured through ICD-10 codes are available (27).

Landmark analysis allowed us to compare the risk of outcomes at a fixed point in time, which reduces the risk of bias and variability at time-zero due to variations in time to diagnosis. The limitation of using a landmark model is that patient characteristics change at a different time during follow-up i.e. an adult that did not receive a diagnosis in 2 years might receive a diagnosis the subsequent year but they will still be under the ‘no diagnosis’ group. The landmark date was chosen as 2 years based on clinical acumen and the time expected for most patients to receive a relevant diagnosis. If a longer period were used, i.e. a 5 year landmark date, then all adults who received a diagnosis would have been correctly identified. However, all adults would have needed to survive up to the 5 year landmark date or be handled using competing risks analysis to be included and adults that died or were lost to follow-up would have to be excluded, reducing sample size (28).

Overall, our findings highlight the burden of breathlessness to the healthcare system. We report novel findings to support the importance of making a timely diagnosis within six months after adults seek healthcare for chronic breathlessness. In individuals where no explanatory diagnosis is recorded, outcomes are better and attention can be focused on interventions to reduce their burden of the symptom (for example, breathing retraining, weight reduction and progressive exercise reconditioning). For individuals who have an underlying cardiorespiratory disease, our data suggest prompt diagnosis may improve outcomes. We therefore propose that the deployment of easy to navigate diagnostic pathways for breathlessness in primary care could achieve better outcomes.

## Data Availability

The study uses data from the Clinical Practice Research Datalink (CPRD). CPRD does not allow the sharing of patient-level data.

## References

1. Sandberg J, Ekström M, Börjesson M, Bergström G, Rosengren A, Angerås O, et al. Underlying contributing conditions to breathlessness among middle-aged individuals in the general population: a cross-sectional study. BMJ Open Respiratory Research. 2020;7(1):e000643.

2. Ferry OR, Huang YC, Masel PJ, Hamilton M, Fong KM, Bowman RV, et al. Diagnostic approach to chronic dyspnoea in adults. Journal of thoracic disease. 2019;11(Suppl 17):S2117.

3. Guo YL, Ampon MR, Poulos LM, Davis SR, Toelle BG, Marks GB, et al. Contribution of obesity to breathlessness in a large nationally representative sample of Australian adults. Respirology. 2023;28(4):350–6.

4. Jagana R, Bartter T, Joshi M. Delay in diagnosis of chronic obstructive pulmonary disease: reasons and solutions. Current Opinion in Pulmonary Medicine. 2015;21(2):121–6.

5. Jones RC, Price D, Ryan D, Sims EJ, von Ziegenweidt J, Mascarenhas L, et al. Opportunities to diagnose chronic obstructive pulmonary disease in routine care in the UK: a retrospective study of a clinical cohort. The Lancet Respiratory Medicine. 2014;2(4):267–76.

6. Cosgrove GP, Bianchi P, Danese S, Lederer DJ. Barriers to timely diagnosis of interstitial lung disease in the real world: the INTENSITY survey. BMC pulmonary medicine. 2018;18:1–9.

7. National Institute for Health and Care Excellence. Breathlessness: Management 2017 [Available from: https://cks.nice.org.uk/breathlessness.

8. NHS England. Adult breathlessness pathway (pre-diagnosis): diagnostic pathway support tool 2023 [Available from: https://www.england.nhs.uk/publication/adult-breathlessness-pathway-pre-diagnosis-diagnostic-pathway-support-tool/.

9. Watson J, Nicholson BD, Hamilton W, Price S. Identifying clinical features in primary care electronic health record studies: methods for codelist development. BMJ open. 2017;7(11):e019637.

10. Herrett E, Gallagher AM, Bhaskaran K, Forbes H, Mathur R, Van Staa T, et al. Data resource profile: clinical practice research datalink (CPRD). International journal of epidemiology. 2015;44(3):827–36.

11. Primary Care Informatics. Read Terms for General Practice 2014 [Available from: https://www.scimp.scot.nhs.uk/better-information/clinical-coding/scimp-guide-to-read-codes.

12. World Health Organization. ICD-10 Version:2019 2019 [Available from: https://icd.who.int/browse10/2019/en.

13. Payne RA, Mendonca SC, Elliott MN, Saunders CL, Edwards DA, Marshall M, et al. Development and validation of the Cambridge Multimorbidity Score. Cmaj. 2020;192(5):E107–E14.

14. CRAN. Package ‘mice’ 2021 [Available from: https://cran.r-project.org/web/packages/mice/mice.pdf.

15. Doe GE, Williams MT, Chantrell S, Steiner MC, Armstrong N, Hutchinson A, et al. Diagnostic delays for breathlessness in primary care: a qualitative study to investigate current care and inform future pathways. British Journal of General Practice. 2023;73(731):e468–e77.

16. Doe G, Clanchy J, Wathall S, Chantrell S, Edwards S, Baxter N, et al. Feasibility study of a multicentre cluster randomised control trial to investigate the clinical and cost-effectiveness of a structured diagnostic pathway in primary care for chronic breathlessness: protocol paper. BMJ open. 2021;11(11):e057362.

17. Herrett E, Gallagher AM, Bhaskaran K, Forbes H, Mathur R, van Staa T, et al. Data Resource Profile: Clinical Practice Research Datalink (CPRD). Int J Epidemiol. 2015;44(3):827–36.

18. Statistics OfN. 2011 census: Population and household estimates for the United Kingdom. 2011.

19. BMA. Quality and outcomes framework (QOF) 2023 [Available from: https://www.bma.org.uk/advice-and-support/gp-practices/funding-and-contracts/quality-and-outcomes-framework-qof.

20. NHS Digital. National Quality and Outcomes Framework Statistics for England 2006/07. National Statistics. 2006.

21. Taggar JS, Coleman T, Lewis S, Szatkowski L. The impact of the Quality and Outcomes Framework (QOF) on the recording of smoking targets in primary care medical records: cross-sectional analyses from The Health Improvement Network (THIN) database. BMC Public Health. 2012;12(1):329.

22. Chen Y, Hayward R, Chew-Graham CA, Hubbard R, Croft P, Sims K, et al. Prognostic value of first-recorded breathlessness for future chronic respiratory and heart disease: a cohort study using a UK national primary care database. British Journal of General Practice. 2020;70(693):e264–e73.

23. Jensen K, Soguero-Ruiz C, Oyvind Mikalsen K, Lindsetmo R-O, Kouskoumvekaki I, Girolami M, et al. Analysis of free text in electronic health records for identification of cancer patient trajectories. Scientific reports. 2017;7(1):46226.

24. Currow DC, Chang S, Ekström M, Hutchinson A, Luckett T, Kochovska S, et al. Health service utilisation associated with chronic breathlessness: random population sample. ERJ open research. 2021;7(4).

25. Müllerová H, Lu C, Li H, Tabberer M. Prevalence and burden of breathlessness in patients with chronic obstructive pulmonary disease managed in primary care. PloS one. 2014;9(1):e85540.

26. Mahler DA, Harver A, Lentine T, Scott JA, Beck K, Schwartzstein RM. Descriptors of breathlessness in cardiorespiratory diseases. American journal of respiratory and critical care medicine. 1996;154(5):1357–63.

27. OpenSAFELY. SystmOne primary care [Available from: https://docs.opensafely.org/data-sources/systmone/.

28. Hopkinson N. Opinion B, editor2017. Available from: https://blogs.bmj.com/bmj/2017/09/11/nick-hopkinson-chronic-breathlessness-syndrome-the-power-of-a-name/#:~:text=The%20agreed%20term%2C%20%E2%80%9Cchronic%20breathlessness,and%20that%20results%20in%20disability%E2%80%9D.

