## Appendix for "Time to diagnosis and long-term outcomes for adults presenting with breathlessness"

**Appendix A Codelists**

**Appendix B Acute breathlessness**

**Appendix C Modelling**

### Appendix A Codelists

**Table A1.** Read codes used to define first-recorded code for breathlessness in CPRD.

| Medcode | Read code | Read term |
| --- | --- | --- |
| 735 | R060D00 | [D] Breathlessness |
| 3092 | R060A00 | [D] Dyspnoea |
| 100954 | R060E00 | [D] Mild wheeze |
| 101037 | R060F00 | [D] Moderate wheeze |
| 11451 | R060200 | [D] Orthopnoea |
| 101073 | R060G00 | [D] Severe wheeze |
| 741 | R060800 | [D] Shortness of breath |
| 101421 | R060H00 | [D] Very severe wheeze |
| 2210 | R060900 | [D] Wheezing |
| 31143 | 1734 | Breathless - at rest |
| 7683 | 1735 | Breathless - lying flat |
| 7932 | 1733 | Breathless - mild exertion |
| 6326 | 1732 | Breathless - moderate exertion |
| 24889 | 173G.00 | Breathless - strenuous exertion |
| 1429 | 173..00 | Breathlessness |
| 21801 | 173Z.00 | Breathlessness NOS |
| 5175 | 173..11 | Breathlessness symptom |
| 60096 | ZR3Q.00 | CLASP shortness of breath score |
| 57903 | 388H.00 | CLASP shortness of breath score |
| 2931 | 1738 | Difficulty breathing |
| 5896 | 173..12 | Dyspnoea - symptom |
| 53771 | 173C.11 | Dyspnoea on exertion |
| 19432 | 173H.00 | MRC Breathlessness Scale: grade 1 |
| 19427 | 173I.00 | MRC Breathlessness Scale: grade 2 |
| 19426 | 173J.00 | MRC Breathlessness Scale: grade 3 |
| 19430 | 173K.00 | MRC Breathlessness Scale: grade 4 |
| 19429 | 173L.00 | MRC Breathlessness Scale: grade 5 |
| 18116 | 173D.00 | Nocturnal dyspnoea |
| 9089 | 1735.11 | Orthopnoea symptom |
| 6434 | 1736 | Paroxysmal nocturnal dyspnoea |
| 22094 | 173F.00 | Short of breath dressing/undressing |
| 4822 | 1739 | Shortness of breath |
| 2575 | 173C.00 | Short of breath on exertion |
| 5349 | 173..13 | Shortness of breath symptom |
| 12474 | 173C.12 | SOBOE |
| 173 | 1737 | Wheezing |

[D] terms are defined in the Read thesaurus as ‘Symptoms, signs and ill-defined conditions’; CLASP, cardiovascular limitations and symptoms profile; MRC, Medical Research Council; NOS, not otherwise specified; O/E, on examination; SOBOE, shortness of breath on exertion.

**Table A2.** Read codes used to identify LRTI.

| Medcode | Readterm |
| --- | --- |
| 99214 | [X]Acute bronchiolitis due to other specified organisms |
| 73100 | [X]Acute bronchitis due to other specified organisms |
| 98257 | [X]Flu+oth respiratory manifestations,'flu virus identified |
| 63776 | [X]Haemophilus influenzae infection, unspecified |
| 97279 | [X]Influenza+other manifestations, virus not identified |
| 97936 | [X]Influenza+other manifestations,influenza virus identified |
| 66397 | [X]Other acute lower respiratory infections |
| 24800 | Acute bacterial bronchitis unspecified |
| 1019 | Acute bronchiolitis |
| 105895 | Acute bronchiolitis due to human metapneumovirus |
| 66228 | Acute bronchiolitis due to other specified organisms |
| 18451 | Acute bronchiolitis due to respiratory syncytial virus |
| 17917 | Acute bronchiolitis NOS |
| 17185 | Acute bronchiolitis with bronchospasm |
| 312 | Acute bronchitis |
| 29669 | Acute bronchitis and bronchiolitis |
| 93153 | Acute bronchitis due to coxsackievirus |
| 65916 | Acute bronchitis due to echovirus |
| 31886 | Acute bronchitis due to mycoplasma pneumoniae |
| 29273 | Acute bronchitis due to parainfluenza virus |
| 48593 | Acute bronchitis due to respiratory syncytial virus |
| 64890 | Acute bronchitis due to rhinovirus |
| 20198 | Acute bronchitis NOS |
| 41137 | Acute bronchitis or bronchiolitis NOS |
| 21145 | Acute croupous bronchitis |
| 69192 | Acute exudative bronchiolitis |
| 21492 | Acute haemophilus influenzae bronchitis |
| 6124 | Acute lower respiratory tract infection |
| 37447 | Acute lower respiratory tract infection |
| 49794 | Acute neisseria catarrhalis bronchitis |
| 9043 | Acute pneumococcal bronchitis |
| 11072 | Acute purulent bronchitis |
| 21113 | Acute respiratory infection NOS |
| 8025 | Acute respiratory infections |
| 43362 | Acute streptococcal bronchitis |
| 11101 | Acute tracheobronchitis |
| 1382 | Acute viral bronchitis unspecified |
| 5978 | Acute wheezy bronchitis |
| 94930 | Avian influenza |
| 63697 | Avian influenza virus nucleic acid detection |
| 68 | Chest infection |
| 30653 | Chest infection - pneumonia organism OS |
| 17359 | Chest infection - unspecified bronchitis |
| 2581 | Chest infection NOS |
| 24316 | Chest infection with infectious disease EC |
| 7714 | Haemophilus influenzae infection |
| 96286 | Human parainfluenza virus detected |
| 556 | Influenza |
| 98102 | Influenza A (H1N1) swine flu |
| 98143 | Influenza A virus H1N1 subtype detected |
| 97062 | Influenza A virus, other or untyped strain detected |
| 105541 | Influenza B nucleic acid detection |
| 96017 | Influenza B virus detected |
| 98129 | Influenza due to Influenza A virus subtype H1N1 |
| 96019 | Influenza H1 virus detected |
| 102918 | Influenza H2 virus detected |
| 96018 | Influenza H3 virus detected |
| 98156 | Influenza H5 virus detected |
| 5947 | Influenza like illness |
| 16388 | Influenza NOS |
| 15774 | Influenza with laryngitis |
| 47472 | Influenza with other manifestations |
| 31363 | Influenza with other manifestations NOS |
| 43625 | Influenza with other respiratory manifestation |
| 29617 | Influenza with pharyngitis |
| 15912 | Influenza with pneumonia |
| 35745 | Influenza with pneumonia NOS |
| 23488 | Influenza with respiratory manifestations NOS |
| 8980 | Influenza-like symptoms |
| 3358 | Lower resp tract infection |
| 23640 | Other specified acute respiratory infections |
| 11849 | Other specified pneumonia or influenza |
| 94130 | Parainfluenza type 1 nucleic acid detection |
| 94858 | Parainfluenza type 2 nucleic acid detection |
| 91123 | Parainfluenza type 3 nucleic acid detection |
| 104614 | Parainfluenza type 4 nucleic acid detection |
| 36675 | Pneumonia due to parainfluenza virus |
| 6094 | Pneumonia or influenza NOS |
| 7074 | Respiratory infection NOS |
| 12573 | Respiratory syncytial virus infection |
| 293 | Respiratory tract infection |
| 55391 | Subacute bronchitis unspecified |

**Table A3.** Read codes used to identify URTI.

| Medcode | Readterm |
| --- | --- |
| 76 | Upper respiratory infection nos |
| 142 | Acute laryngitis |
| 310 | Throat infection - pharyngitis |
| 368 | Common cold |
| 407 | Acute pharyngitis nos |
| 893 | Acute pharyngitis |
| 1765 | Streptococcal sore throat |
| 2637 | Upper respiratory tract infection nos |
| 3260 | Acute nasopharyngitis |
| 4221 | Recurrent upper respiratory tract infection |
| 4718 | Pharyngolaryngitis |
| 4868 | Acute viral pharyngitis |
| 4902 | Streptococcal pharyngitis |
| 5115 | Acute viral laryngitis unspecified |
| 5553 | Has a sore throat |
| 5755 | Sore throat symptom |
| 6014 | Sore throat nos |
| 6294 | Acute upper respiratory tract infection |
| 6421 | Viral upper respiratory tract infection nos |
| 6466 | Viral sore throat nos |
| 6620 | Febrile cold |
| 8950 | Feverish cold |
| 9093 | Pyrexial cold |
| 10093 | Tracheopharyngitis |
| 10765 | Acute catarrhal laryngitis |
| 11499 | Throat infection - tonsillitis |
| 12489 | Persistent sore throat |
| 15287 | Sore throat symptom nos |
| 15628 | Other upper respiratory infections of multiple sites |
| 15774 | Influenza with laryngitis |
| 16120 | Acute laryngitis and tracheitis nos |
| 16184 | Streptococcal sore throat with scarlatina nos |
| 16217 | Streptococcal sore throat nos |
| 16718 | Streptococcal laryngitis |
| 17899 | Acute bacterial pharyngitis |
| 18908 | Acute laryngopharyngitis |
| 21486 | Acute ulcerative pharyngitis |
| 22720 | Acute laryngitis nos |
| 24708 | Acute phlegmonous pharyngitis |
| 26010 | Other acute upper respiratory infections |
| 29589 | Acute staphylococcal pharyngitis |
| 29617 | Influenza with pharyngitis |
| 31501 | Acute phlegmonous laryngitis |
| 41324 | Acute laryngitis and tracheitis |
| 43317 | Acute haemophilus influenzae laryngitis |
| 51562 | Acute suppurative laryngitis |
| 52756 | Acute bacterial laryngitis unspecified |
| 53055 | [X]acute upper respiratory infections |
| 53395 | Acute bacterial pharyngitis nos |
| 54777 | Streptococcal sore throat and scarlatina |
| 62885 | Acute ulcerative laryngitis |
| 92428 | Acute pneumococcal pharyngitis |
| 93964 | [x]Acute pharyngitis due to other specified organisms |

**Table A4.** Read codes used to identify pneumonia.

| Medcode | Readterm |
| --- | --- |
| 572 | Pneumonia due to unspecified organism |
| 886 | Bronchopneumonia due to unspecified organism |
| 1576 | Pneumonia due to Mycoplasma pneumoniae |
| 5612 | Pneumonia due to staphylococcus |
| 6094 | Pneumonia or influenza NOS |
| 9639 | Lobar pneumonia due to unspecified organism |
| 10086 | Pneumonia and influenza |
| 11849 | Other specified pneumonia or influenza |
| 12061 | Pneumonia - Legionella |
| 12423 | Pneumonia due to Streptococcus |
| 14976 | Viral pneumonia NOS |
| 23095 | Bacterial pneumonia NOS |
| 23546 | Pneumonia due to Klebsiella pneumoniae |
| 23726 | Pneumonia with varicella |
| 25694 | Pneumonia due to other specified organisms |
| 27519 | Pneumonia with pneumocystis carinii |
| 30437 | Pneumonia with whooping cough |
| 30591 | Pneumonia due to Pseudomonas |
| 30653 | Chest infection - pneumonia organism OS |
| 31269 | Pneumonia due to respiratory syncytial virus |
| 33478 | Viral pneumonia NEC |
| 34251 | Pneumonia due to specified organism NOS |
| 34274 | Pneumonia with aspergillosis |
| 35082 | Pneumonia with pertussis |
| 35745 | Influenza with pneumonia NOS |
| 36675 | Pneumonia due to parainfluenza virus |
| 37881 | Pneumonia due to haemophilus influenzae |
| 40299 | Pneumonia - candidal |
| 40498 | Pneumonia with infectious diseases EC |
| 41034 | Pneumonia with measles |
| 43286 | Pneumonia with cytomegalic inclusion disease |
| 43884 | Pneumonia due to bacteria NOS |
| 45425 | Pneumonia due to proteus |
| 48804 | Pneumonia due to haemophilus influenzae |
| 49398 | Pneumonia with typhoid fever |
| 50867 | Pneumonia due to other specified bacteria |
| 52071 | Pneumonia with candidiasis |
| 52384 | Pneumonia due to other aerobic gram-negative bacteria |
| 53947 | [X]Pneumonia in viral diseases classified elsewhere |
| 53969 | Pneumonia with systemic mycosis NOS |
| 60482 | Pneumonia with Q-fever |
| 61623 | Pneumonia with actinomycosis |
| 62623 | Pneumonia with ornithosis |
| 63858 | Pneumonia due to streptococcus, group B |
| 66362 | Pneumonia with infectious diseases EC NOS |
| 67836 | Pneumonia due to adenovirus |
| 67901 | Pneumonia with nocardiasis |
| 69782 | Pneumonia with other infectious diseases EC |
| 70559 | Pneumonia with other infectious diseases EC NOS |
| 72182 | Pneumonia with salmonellosis |
| 73735 | Pneumonia due to pleuropneumonia like organisms |
| 98381 | [X]Pneumonia due to other specified infectious organisms |
| 98782 | Pneumonia with toxoplasmosis |
| 103404 | Pneumonia with coccidioidomycosis |
| 106031 | [X]Mycoplasma pneumoniae [PPLO]cause/dis classifd/oth chaptr |
| 106300 | Pneumonia due to Human metapneumovirus |
| 106908 | Pneumonia with tularaemia |
| 110440 | Pneumonia with other systemic mycoses |
| 111027 | [X]Pneumonia in bacterial diseases classified elsewhere |
| 111655 | [X]Pneumonia in other diseases classified elsewhere |
| 114636 | [X]Pneumonia due to other aerobic gram-negative bacteria |
| 22795 | Chest infection - other bacterial pneumonia |
| 13573 | Influenza with bronchopneumonia |
| 5202 | Viral pneumonia |
| 57667 | Gangrenous pneumonia |
| 104121 | Community acquired pneumonia |
| 32172 | Postmeasles pneumonia |
| 28634 | Other bacterial pneumonia |
| 62632 | Influenza with pneumonia, influenza virus identified |
| 52520 | [X]Other viral pneumonia |
| 63763 | [X]Other bacterial pneumonia |
| 53753 | [X]Other pneumonia, organism unspecified |
| 5324 | Atypical pneumonia |
| 17025 | Chlamydial pneumonia |
| 47973 | Herpes simplex pneumonia |
| 29457 | Chest infection - influenza with pneumonia |
| 29166 | Chest infection - pneumococcal pneumonia |
| 16287 | Chest infection - unspecified bronchopneumonia |
| 9389 | Chest infection - viral pneumonia |
| 60299 | E.coli pneumonia |
| 1849 | Lobar (pneumococcal) pneumonia |
| 26287 | Klebsiella pneumoniae/cause/disease classifd/oth chapters |
| 15912 | Influenza with pneumonia |
| 19400 | Chest infection - pneumonia due to unspecified organism |

**Read and ICD-10 diagnoses codes**

Read codes and ICD-10 codes were established for the underlying causes of breathlessness. Diagnoses can be made in both primary care and secondary care, hence, were applied in CPRD and HES. Diagnoses have been split into three categories: cardiac, pulmonary and other.

**Table A5.** Number of Read and ICD-10 codes used to establish diagnoses.

| Diagnosis | Read codes | ICD-10 codes |
| --- | --- | --- |
| Cardiac | 307 | 160 |
| Heart failure | 28 | 34 |
| Left ventricular hypertrophy | 2 | 1 |
| Right ventricular hypertrophy | 1 | 0 |
| Coronary heart disease | 59 | 10 |
| Cardiomyopathy | 50 | 14 |
| Valvular disease | 50 | 49 |
| Cardiac arrhythmias / atrial fibrillation | 37 | 20 |
| Pericardial disease | 20 | 14 |
| Congenital heart disease | 60 | 18 |
| Pulmonary | 137 | 116 |
| COPD | 22 | 16 |
| Asthma | 36 | 21 |
| Interstitial lung disease | 15 | 23 |
| Lung cancer | 24 | 21 |
| Pleural effusion | 3 | 2 |
| Bronchiectasis | 6 | 4 |
| Thromboembolic disease | 17 | 1 |
| Pulmonary hypertension | 3 | 15 |
| Chest wall deformities | 5 | 2 |
| Neuromuscular disorders | 6 | 11 |
| Other | 587 | 6 |
| Anaemia | 191 | 21 |
| Anxiety | 305 | 6 |
| Depression | 73 | 11 |
| Obesity | 16 | 9 |
| Dysfunctional breathing | 2 | 9 |

**Table A6.** Linked publications/sources used to establish Read coded diagnoses.

| Cardiac |  |
| --- | --- |
| Heart failure | Real-world presentation with heart failure in primary care: do patients selected to follow diagnostic and management guidelines have better outcomes? - Alex Bottle [Open Heart 2018] |
| Left ventricular hypertrophy | CPRD code browser search terms: *ventricular hypertrophy* |
| Right ventricular hypertrophy | CPRD code browser search terms: *ventricular hypertrophy* |
| Coronary heart disease | Effect of financial incentives on incentivised and non-incentivised clinical activities: longitudinal analysis of data from the UK Quality and Outcomes Framework - Tim Doran [British Medical Journal 2011] |
| Cardiomyopathy | Matthews, A and Bhaskaran, K (2017). *Clinical code list - Cardiomyopathy.* [Data Collection]. London School of Hygiene & Tropical Medicine, London, United Kingdom. https://doi.org/10.17037/DATA.195. |
| Valvular disease | Matthews, A and Bhaskaran, K (2018). *Clinical code list - Valvular heart disease.* [Data Collection]. London School of Hygiene & Tropical Medicine, London, United Kingdom. https://doi.org/10.17037/DATA.00000732. |
| Cardiac arrhythmias / Atrial fibrillation | Discontinuation of non-Vitamin K antagonist oral anticoagulants in patients with non-valvular atrial fibrillation: a population-based cohort study using primary care data from The Health Improvement Network in the UK - Ana Ruigómez [BMJ Open 2019] |
|  | Codes for detection cardiac arrhythmias - http://links.lww.com/EDE/A145 |
| Pericardial disease | Matthews, A and Bhaskaran, K (2018). *Clinical code list - Pericarditis.* [Data Collection]. London School of Hygiene & Tropical Medicine, London, United Kingdom. https://doi.org/10.17037/DATA.00000731. |
| Congenital heart disease | Determining the predictive value of Read codes to identify congenital cardiac malformations in the UK Clinical Practice Research Datalink - Tarek A. Hammad [Pharmacoepidemiology and Drug Safety - 2013] |
| Pulmonary |  |
| COPD | Validation of chronic obstructive pulmonary disease recording in the Clinical Practice Research Datalink (CPRD-GOLD) - Jenni Quint [BMJ Open 2014] |
|  | Changing causes of death for patients with chronic respiratory disease in England, 2005-2015 - Jenni Quint [Thorax 2019] |
| Asthma | Prognostic value of first-recorded breathlessness for future chronic respiratory and heart disease: a cohort study using a UK national primary care database - Ying Chen [British Journal of Gen Practice 2020] |
| Interstitial lung disease | Interstitial lung disease is a risk factor for ischaemic heart disease and myocardial infarction - Lorna E. Clarson [Heart 2020] |
| Lung cancer | Can analyses of electronic patient records be independently and externally validated? Study 2: the Effect of Beta-Adrenoceptor Blocker Therapy on Cancer Survival; a Retrospective Cohort Study - David A. Springate [BMJ Open 2015] |
| Pleural effusion | Long-term trends of use of health service among heart failure patients - Alex Bottle [European Heart Journal - Quality of Care and Clinical Outcomes 2018] |
| Bronchiectasis | ERS Supplementary Material - https://erj.ersjournals.com/content/suppl/2015/11/05/13993003.01033-2015.DC2 |
| Thromboembolic disease | Discontinuation of non-Vitamin K antagonist oral anticoagulants in patients with non-valvular atrial fibrillation: a population-based cohort study using primary care data from The Health Improvement Network in the UK - Ana Ruigómez [BMJ Open 2019] |
| Pulmonary hypertension | CPRD code browser search terms: *pulmonary hypertension* |
| Chest wall deformities | CPRD code browser search terms: *kyphoscoliosis*, *chest wall deform* |
| Neuromuscular disorders | CPRD code browser search terms: *myasthenia gravis*, *amyotrophic lateral sclerosis*, *amyotrophic lateral sclerosis* |
| Other |  |
| Anaemia | Malhotra, A (2020). Clinical code list - Anaemia. [Data Collection]. London School of Hygiene & Tropical Medicine, London, United Kingdom. https://doi.org/10.17037/DATA.00001920. |
| Anxiety | Suicide risk linked with clinical consultation frequency, psychiatric diagnoses and psychotropic medication prescribing in a national study of primary-care patients - K Winfuhr [Psychol Med 2016 Dec;46(16):3407-3417] |
| Depression | Long-term trends of use of health service among heart failure patients - Alex Bottle [European Heart Journal - Quality of Care and Clinical Outcomes 2018] |
| Obesity | Cardiovascular Risk and Risk Factor Management in Type 2 Diabetes Mellitus - Alison Wright [Circulation 2019 Jun 11;139(24):2742-2753] |
| Dysfunctional breathing | CPRD code browser search terms: *hyperventilation* |

**Table A7.** Linked publications/sources used to establish ICD-10 coded diagnoses.

| Cardiac | |
| --- | --- |
| Heart failure | <https://www.icd10data.com/ICD10CM/Codes/I00-I99/I30-I52/I50-> |
| Left ventricular hypertrophy | https://www.icd10data.com/ICD10CM/Codes/I00-I99/I30-I5A/I51-/I51.7#:~:text=Billable%2FSpecific%20Code-,I51.,ICD%2D10%2DCM%20I51. |
| Right ventricular hypertrophy* | None |
| Coronary heart disease | <https://www.icd10data.com/ICD10CM/Codes/I00-I99/I20-I25> |
| Cardiomyopathy | https://www.icd10data.com/ICD10CM/Codes/I00-I99/I30-I52/I42-/I42 |
| Valvular disease | Matthews, A and Bhaskaran, K (2018). *Clinical code list - Valvular heart disease.* [Data Collection]. London School of Hygiene & Tropical Medicine, London, United Kingdom. https://doi.org/10.17037/DATA.00000732. |
| Cardiac arrhythmias / Atrial fibrillation | https://icd.codes/icd10cm/I48 |
|  | <https://www.icd10data.com/ICD10CM/Codes/I00-I99/I30-I52/I49-> |
| Pericardial disease | Matthews, A and Bhaskaran, K (2018). *Clinical code list - Pericarditis.* [Data Collection]. London School of Hygiene & Tropical Medicine, London, United Kingdom. https://doi.org/10.17037/DATA.00000731. |
| Congenital heart disease | https://www.icd10data.com/ICD10CM/Codes/Q00-Q99/Q20-Q28/Q24-/Q24.9 |
| Pulmonary | |
| COPD | <https://www.copdfoundation.org/pdfs/ICD%20Reference%20Codes.pdf> |
| Asthma | https://www.copdfoundation.org/pdfs/ICD%20Reference%20Codes.pdf |
| Interstitial lung disease | <https://www.icd10data.com/ICD10CM/Codes/J00-J99/J80-J84/J84-/J84.9#:~:text=2021%20ICD%2D10%2DCM%20Diagnosis,9%3A%20Interstitial%>  20pulmonary%20disease%2C%20unspecified[https://www.icd10data.com/ICD10CM/Codes/J00-J99/J80-J84/J84-/J84.9](https://www.icd10data.com/ICD10CM/Codes/J00-J99/J80-J84/J84-/J84.9#:~:text=2021%20ICD%2D10%2DCM%20Diagnosis,9%3A%20Interstitial%20pulmonary%20disease%2C%20unspecified) |
|  | https://www.thoracic.org/about/newsroom/newsletters/coding-and-billing/2015/september/icd-10-cm-coding.php |
| Lung cancer | <https://www.icd10data.com/ICD10CM/Codes/C00-D49/C30-C39/C34-/C34.90> |
| Pleural effusion | <https://www.icd10data.com/ICD10CM/Codes/J00-J99/J90-J94/J90-/J90#:~:text=Pleural%20effusion%2C%20not%20elsewhere%20classified,-2016%202017%202018&text=J90%20is%20a%20billable%2Fspecific,a%>  20diagnosis%20for%20reimbursement%20purposes. |
| Bronchiectasis | https://www.icd10data.com/search?s=bronchiectasis |
| Thromboembolic disease | <https://www.icd10data.com/ICD10CM/Codes/I00-I99/I26-I28/I27-/I27.82> |
| Pulmonary hypertension | <https://www.outsourcestrategies.com/resources/learn-the-correct-diagnosis-codes-for-pulmonary-hypertension-ph.html> |
| Chest wall deformities | <https://www.icd10data.com/ICD10CM/Codes/Q00-Q99/Q65-Q79/Q67-/Q67.8> |
|  | <https://www.icd10data.com/search?s=M41> |
| Neuromuscular disorders | <https://www.slideshare.net/osimos/documenting-myasthenia-gravis-with-icd-10-codes> |
|  | <https://www.icd10data.com/ICD10CM/Codes/G00-G99/G10-G14/G12-/G12.21> |
| Other | |
| Anaemia | <https://www.icd10data.com/ICD10CM/Codes/D50-D89/D60-D64/D64-/D64.9> |
| Anxiety | <https://www.icd10data.com/ICD10CM/Codes/F01-F99/F40-F48/F41-/F41.9> |
| Depression | <https://www.icd10data.com/ICD10CM/Codes/F01-F99/F30-F39/F33-> |
| Obesity | <https://www.icd10data.com/ICD10CM/Codes/E00-E89/E65-E68> |
| Dysfunctional breathing | <https://www.icd10data.com/ICD10CM/Codes/R00-R99/R00-R09/R06-> |

*No ICD-10 codes were found for right ventricular hypertrophy.

**Read coded diagnoses (medcodes)**

Read codes and Read terms were compiled to define diagnoses in primary care for each cardiac, pulmonary and other condition. The CPRD codes diagnoses using medcodes so Read codes were translated and applied as appropriate to the Clinical file.

**Table A8.** List of medcodes applied in CPRD to establish diagnoses.

| Cardiac | |
| --- | --- |
| Heart failure | 884 5255 10154 104275 398 1223 2062 2906 4024 9524 10079 11424 17278 52127 57987 62718 66306 21837 23707 27884 27964 32671 72668 94870 96799 101137 101138 106897 |
| Left ventricular hypertrophy | 5942 8966 |
| Right ventricular hypertrophy | 12550 |
| Coronary heart disease | 240 27951 15661 24783 36523 7347 4656 39655 1431 19655 17307 34328 18118 11983 54251 20416 39449 1792 9413 9276 39693 27977 1430 20095 18125 29902 12986 11048 36854 25842 66388 54535 7696 1414 32450 9555 26863 12804 28554 28138 5413 3999 5254 1655 1344 36609 7320 29421 34633 24540 23078 35713 15754 18889 22383 1676 52517 68401 47637 |
| Cardiomyopathy | 39423 68677 23452 72175 44272 62404 7320 98167 72409 104373 107257 71848 55416 3204 8010 68685 57306 5141 68766 41488 21852 3499 7535 101015 104658 40834 100966 64673 55850 70855 27683 64837 105651 98020 97780 102955 104529 42043 9402 58938 22993 10415 34437 49787 73153 70648 92266 97617 98634 40095 |
| Valvular disease | 32211 7963 63960 47887 10187 999 58810 1005 10964 9591 6886 8636 58734 6843 51879 22837 44488 44328 28662 49355 17596 33262 33907 94872 11878 40949 9450 561 61651 54088 105626 15640 15496 38299 6077 34932 65000 96754 60266 21980 93114 93113 42128 9286 72306 49551 97738 35372 52271 5743 |
| Cardiac arrhythmias / Atrial fibrillation | 96076 96277 2212 1664 1757 1268 35127 23437 6503 53893 4044 70366 4374 4827 25583 5484 41916 7457 2249 2579 19979 3909 4802 9023 29654 31809 27413 426 7827 4421 23494 8651 31690 31133 1535 20011 21955 |
| Pericardial disease | 96444 94380 35119 50720 96101 20157 65807 18293 105192 108258 11920 16803 9113 65897 96449 8411 44376 72628 107662 18877 |
| Congenital heart disease | 46 3255 2727 3625 7474 3301 4864 6886 2670 9401 3300 247 22778 23692 2816 3774 20062 5621 31112 34067 8279 11127 24789 25481 39992 3862 7262 10505 16539 3863 18395 18982 20772 30725 33919 42132 89256 90486 9232 18785 21851 24533 32508 34668 38968 41371 43560 44767 44896 45187 45452 46176 50362 51941 54487 54488 54772 64953 68784 69940 |
| Pulmonary | |
| COPD | 794 998 1001 5710 9876 10802 10863 10980 14798 23492 26306 33450 37247 44525 93568 12166 104608 65733 67040 60188 46578 16410 |
| Asthma | 5267 2290 1208 15248 7731 93353 4892 1555 106805 7146 232 14777 185 47684 18323 78 12987 4442 25796 8335 21232 5798 58196 6707 109958 16070 29325 40823 27926 3665 45782 39478 45073 233 4606 5627 |
| Interstitial lung disease | 3859 3865 4910 5519 6051 6837 8317 22905 28229 33980 58841 63174 103472 103559 103753 |
| Lung cancer | 13243 15221 103946 37810 12870 17391 33444 21698 10358 31700 25886 44169 31268 41523 39923 54134 31188 18678 12582 42566 36371 7484 38961 3903 |
| Pleural effusion | 9559 947 52850 |
| Bronchiectasis | 2195 32679 20364 15693 41491 56427 |
| Thromboembolic disease | 1266 96209 18121 4717 9701 67006 68438 7174 49269 112578 109337 73569 101944 97367 44404 98639 16976 |
| Pulmonary hypertension | 102444 34065 245 |
| Chest wall deformities | 17068 14790 714 28717 11620 |
| Neuromuscular disorders | 27515 66740 55634 5655 36433 104026 |
| Other | |
| Anaemia | 16052 99222 65351 69061 27771 8054 57897 31550 56348 2054 35092 47952 60186 100576 23875 1668 539 57397 53052 15314 56208 69964 56973 34953 104454 58136 24870 62257 10817 1771 12176 53799 57114 37539 32715 51489 15439 11961 72276 33420 37320 31306 48145 23519 53846 19951 50495 15658 45151 795 44527 63936 34754 56756 107820 106993 2452 22890 69275 47225 3326 69379 41699 18137 99917 43166 57954 51169 16929 4475 33634 105439 31370 64601 3981 67088 99494 739 30637 15936 9537 93872 55561 105985 882 29601 10506 45929 7841 58695 70128 26327 39944 39876 43330 31205 94387 21723 16108 19130 25394 66137 103252 2482 56114 99308 19383 94528 18631 71840 94214 100388 15422 14698 8119 797 1702 4670 70762 44913 28768 49182 53422 3818 2743 57298 31248 31734 69027 2464 22531 33708 98709 92106 5271 55481 38327 104740 103151 40750 41142 21127 43825 59103 94921 42117 22715 19574 4839 34934 39967 68087 31040 53783 57859 72721 48338 6816 55370 2813 27726 64625 37082 104812 31270 15633 32953 102848 32937 29486 36961 44420 104843 49451 21119 35160 25876 3265 33278 4080 36634 70835 43367 6028 39456 57575 57274 66239 4858 110176 71808 |
| Anxiety | 9686 5249 636 6939 4069 462 4659 655 1758 4634 4534 2188 41572 3438 44739 23598 34696 16484 33702 15431 46399 4143 4269 4775 43302 23490 29322 24525 23354 4105 24638 40066 1907 2300 3076 16638 9944 12838 16199 31957 18603 28106 28938 1723 31672 4167 1510 2366 10390 6071 14729 38543 3208 5678 2030 47365 15566 3361 1582 7235 5305 966 42000 15321 56941 3685 39518 72171 43050 14780 791 4199 44212 48561 56800 15035 38134 32034 15483 47809 20053 23413 41615 34664 29448 15292 15284 30961 15034 29461 15959 15224 31422 4963 23774 37695 2871 15371 10158 3869 15939 71437 68379 20109 44547 55781 73547 89237 62400 10001 96391 5067 40311 53766 191 9999 45205 54373 276 11940 42737 43550 15551 38640 20245 24847 29707 23869 26138 28129 20802 27742 2826 2775 10734 24212 6221 23327 54658 62193 67304 28302 56924 58013 48588 15665 16415 6075 35914 66398 45603 41455 53362 32387 35632 7716 27390 19921 23462 37669 50106 42410 20773 15220 23808 9386 2571 16729 14890 11602 42788 9785 67965 18248 11280 12635 12508 27685 34064 7222 67898 5385 8205 6408 4081 10344 962 35825 50191 11913 7749 44321 24066 28167 23838 25638 5304 20634 22019 24251 21836 18399 38809 22721 11098 11607 36374 10535 70779 21559 7813 4171 32182 11336 36228 21197 41235 101785 101725 21753 31515 22136 40994 39826 35311 52161 27588 56141 39747 34978 12147 18801 56966 11354 27633 50121 88758 64166 46567 39919 16988 68259 48906 18049 24439 23704 59682 57877 34735 7537 12626 30680 24264 10870 66806 41038 44269 32632 63259 50793 12715 43316 16714 53122 47698 53737 12830 17081 16560 93067 30179 54382 38521 29329 36040 12453 100086 62002 46925 20906 16679 36009 47570 48671 19242 28090 16561 9656 9265 44331 100116 47367 11339 61753 90597 44586 49628 21431 17687 18032 24351 29907 61430 7999 99609 |
| Depression | 6546 6950 595 34390 16506 15155 15219 32159 43324 57409 7011 15099 6932 35671 29342 14709 25697 24171 56273 55384 6482 25563 3702 17385 27491 9183 17770 1055 655 4639 9055 18510 7604 11717 9211 9667 41989 22806 59386 12099 24117 52678 24112 28863 10667 98252 98414 98417 101054 101153 103677 6854 10720 56609 2970 543 3291 5987 3292 8851 19696 8902 23731 28677 16861 37764 22116 47731 44300 36616 42857 26374 35320 |
| Obesity | 66406 430 38059 8854 22695 25968 103574 11401 52782 69757 10728 13278 22556 108694 108610 108478 |
| Dysfunctional breathing | 2100 20053 |

**ICD-10 coded diagnoses**

**Table A9.** List of ICD-10 codes applied in HES to establish diagnoses.

| Cardiac | |
| --- | --- |
| Heart failure | I11 I11.0 I11.9 I13.0 I13.2 I50 I50.1 I50.2 I50.20 I50.21 I50.22 I50.23 I50.3 I50.30 I50.31 I50.32 I50.33 I50.4 I50.40 I50.41 I50.42 I50.43 I50.8 I50.81 I50.810 I50.811 I50.812 I50.813 I50.814 I50.82 I50.83 I50.84 I50.89 I50.9 |
| Left ventricular hypertrophy | I51.7 |
| Right ventricular hypertrophy | None |
| Coronary heart disease | I20 I20.1 I20.8 I20.9 I24 I24.0 I24.1 I24.8 I24.9 I25 |
| Cardiomyopathy | I40.0 I40.1 I41 I42 I42.0 I42.1 I42.2 I42.3 I42.4 I42.5 I42.6 I42.7 I42.8 I42.9 |
| Valvular disease | I05 I05.0 I05.1 I05.2 I05.8 I05.9 I06 I06.0 I06.1 I06.2 I06.8 I06.9 I07 I07.0 I07.1 I07.2 I07.8 I07.9 I08 I08.0 I08.1 I08.2 I08.3 I08.8 I08.9 I34 I34.0 I34.1 I34.2 I34.8 I34.9 I35 I35.0 I35.1 I35.2 I35.8 I35.9 I36 I36.0 I36.1 I36.2 I36.8 I36.9 I37 I37.0 I37.1 I37.2 I37.8 I37.9 |
| Cardiac arrhythmias / Atrial fibrillation | I48 I48.0 I48.1 I48.2 I48.3 I48.4 I48.9 I49 I49.0 I49.01 I49.02 I49.1 I49.2 I49.3 I49.4 I49.40 I49.49 I49.5 I49.8 I49.9 |
| Pericardial disease | I30.1 I30.8 I31 I31.0 I31.1 I31.2 I31.3 I31.4 I31.8 I31.9 I32 I32.0 I32.1 I32.8 |
| Congenital heart disease | Q24 Q24.0  Q24.2  Q24.4  Q24.5  Q24.6  Q24.8  Q24.9  Q25  Q25.0  Q25.1  Q25.2  Q25.21  Q25.29  Q25.3  Q25.4  Q25.40  Q25.41 |
| Pulmonary | |
| COPD | J40 J41 J41.0 J41.1 J41.8 J42 J43 J43.0 J43.1 J43.2 J43.8 J43.9 J44 J44.0 J44.1 J44.9 |
| Asthma | J45 J45.2 J45.20 J45.21 J45.22 J45.3 J45.30 J45.31 J45.32 J45.4 J45.40 J45.41 J45.42 J45.5 J45.50 J45.51 J45.52 J45.9 J45.90 J45.901 J45.902 |
| Interstitial lung disease | J84.01 J84.02 J84.03 J48.1 J84.111 J84.112 J84.113 J84.114 J84.115 J84.116 J84.117 J84.2 J84.8 J84.81 J84.82 J84.83 J84.84 J84.841 J84.842 J84.843 J84.848 J84.89 J84.9 |
| Lung cancer | C34.2 C34.3 C34.30 C34.31 C34.32 C34.8 C34.80 C34.81 C34.82 C34.9 C34.90 C34.91 C34.92 C37 C38 C38.0 C38.1 C38.2 C38.3 C38.4 C38.8 |
| Pleural effusion | J90 J91 |
| Bronchiectasis | J47 J47.0 J47.1 J47.9 |
| Thromboembolic disease | I27.82 |
| Pulmonary hypertension | I27 I27.0 I27.1 I27.2 I27.20 I27.21 I27.22 I27.23 I27.24 I27.29 I27.8 I27.81 I27.83 I27.89 I27.9 |
| Chest wall deformities | Q67.8 41.9 |
| Neuromuscular disorders | G70 G70.00 G70.01 G70.1 G70.2 G70.8 G70.80 G70.81 G70.89 G70.9 G12.21 |
| Other | |
| Anaemia | D63.8 D64 D64.0 D64.1 D64.2 D64.3 D64.4 D64.8 D64.81 D64.89 D64.9 D65 D66 D67 D68 D68.0 D68.1 D68.2 D68.3 D68.31 D68.311 |
| Anxiety | F41 F41.0 F41.1 F41.3 F41.8 F41.9 |
| Depression | F33 F33.0 F33.1 F33.2 F33.3 F33.4 F33.40 F33.41 F33.42 F33.8 F33.9 |
| Obesity | E65 E66 E66.0 E66.01 E66.09 E66.1 E66.2 E66.8 E66.9 |
| Dysfunctional breathing | R06 R06.1 R06.2 R06.3 R06.4 R06.5 R06.8 R06.89 R06.9 |

### Appendix B Baseline characteristics

**Distinguishing an acute presentation of breathlessness**

Patients who received a code for an acute respiratory infection on the same date as breathlessness were classified as having an acute presentation with breathlessness. An acute respiratory infection was defined as having a recorded code for either an upper respiratory tract infection (URTI), lower respiratory tract infection (LRTI) or pneumonia. Medcodes used to define acute respiratory infections can be found above in Appendix A. Overall, 2495 patients were identified as presenting with acute breathlessness. Patient demographics at first-recorded code for breathlessness can be seen below, as well as long-term outcomes of unplanned hospital admission and all-cause mortality.

**Table B1.** Patient characteristics for adults with acute breathlessness.

|  | LRTI | Pneumonia | URTI |
| --- | --- | --- | --- |
| No. of patients, (%) | 1469 (58.88) | 139 (5.57) | 887 (35.55) |
| Gender, n (%) |  |  |  |
| Male | 630 (42.89) | 61 (43.88) | 355 (40.02) |
| Female | 839 (57.11) | 78 (56.12) | 532 (59.98) |
| Mean ± SD, Age (years) | 51 ± 16 | 56 ± 16 | 45 ± 15 |
| Ethnicity, n (%) |  |  |  |
| Black | 14 (0.95) | 1 (0.72) | 13 (1.47) |
| Mixed/Other | 14 (0.95) | 2 (1.44) | 9 (1.01) |
| South Asian | 18 (1.23) | 0 (0.00) | 38 (4.28) |
| White | 1287 (87.61) | 132 (94.96) | 730 (82.30) |
| Unknown/Missing | 136 (9.26) | 4 (2.88) | 97 (10.94) |
| Mean ± SD, BMI (kg/mass) | 35.42 ± 6.18 | 30.54 ± 7.38 | 33.33 ± 5.60 |
| Smoking Status, n (%) |  |  |  |
| Current smoker | 407 (27.71) | 23 (16.55) | 198 (22.32) |
| Non-smoker | 463 (31.52) | 52 (37.41) | 333 (37.54) |
| Ex-smoker | 386 (26.28) | 39 (28.06) | 210 (23.68) |
| Data Not Entered | 213 (14.50) | 25 (17.99) | 146 (16.46) |
| Deprivation, n (%) |  |  |  |
| 1 (least) | 295 (20.08) | 30 (21.58) | 213 (24.01) |
| 2 | 273 (18.58) | 33 (23.74) | 197 (22.21) |
| 3 | 396 (26.96) | 31 (22.30) | 230 (25.93) |
| 4 | 264 (17.97) | 21 (15.11) | 125 (14.09) |
| 5 (most) | 241 (16.41) | 24 (17.27) | 122 (13.75) |
| Comorbidities, n (%) |  |  |  |
| 0 | 331 (22.53) | 26 (18.71) | 209 (23.56) |
| 1 | 364 (24.78) | 35 (25.18) | 245 (27.62) |
| ≥2 | 774 (52.69) | 78 (56.11) | 433 (48.82) |

**Table B2.** Long-term outcomes of unplanned hospital admission and all-cause mortality for adults with acute breathlessness.

|  | LRTI | Pneumonia | URTI |
| --- | --- | --- | --- |
| No. of patients, (%) | 1469 (58.88) | 139 (5.57) | 887 (35.55) |
| Unplanned admission | 1110 (75.56) | 110 (79.14) | 610 (68.77) |
| Died | 132 (8.99) | 22 (15.83) | 34 (3.83) |

**Pre-existing conditions for adults presenting with chronic breathlessness**

As part of the eligibility criteria, only adults with no record of COPD, heart failure, ILD, or asthma (within the last 10 years) were included in the study cohort. However, other pre-existing conditions could be present that were later investigated as a diagnosis. Pre-existing conditions at first-recorded code for breathlessness were investigated.

**Table B3.** Number of pre-existing conditions at index.

|  | All | 1 pre-existing | 2 pre-existing |
| --- | --- | --- | --- |
| Number of patients, n (%) | 37,469 (36.96) | 35,554 (94.89) | 1,915 (5.11) |
| Pre-existing condition, n (%) |  |  |  |
| Cardiorespiratory | 11,085 (29.58) | 11,073 (31.14) | 12 (0.63) |
| Other | 26,217 (69.97) | 24,314 (68.39) | 1,903 (99.37) |
| Other  (Dysfunctional breathing) | 167 (0.45) | 167 (0.47) | 0 (0.00) |

**Table B4.** Type of pre-existing condition present at index.

|  | 1 pre-existing | 2 pre-existing |
| --- | --- | --- |
| Number of patients | 35,554 (94.89) | 1,915 (5.11) |
| Anaemia | 4,194 (11.80) | 0 (0.00) |
| Anxiety | 10,024 (28.19) | 1,903 (99.37) |
| Cardiac arrhythmias / AF | 3,425 (9.63) | 0 (0.00) |
| Bronchiectasis | 358 (1.01) | 0 (0.00) |
| Cardiomyopathy | 109 (0.31) | 1 (0.05) |
| Chest wall deformities | 191 (0.54) | 0 (0.00) |
| Congenital heart disease | 343 (0.96) | 11 (0.57) |
| Coronary heart disease | 5,019 (14.12) | 1 (0.05) |
| Depression | 6,013 (16.91) | 1,895 (98.96) |
| Dysfunctional breathing | 167 (0.47) | 8 (0.42) |
| Lung cancer | 65 (0.18) | 0 (0.00) |
| Left ventricular hypertrophy | 62 (0.17) | 0 (0.00) |
| Neuromuscular disorders | 44 (0.12) | 0 (0.00) |
| Obesity | 4,083 (11.48) | 0 (0.00) |
| Pericardial disease | 32 (0.09) | 0 (0.00) |
| Pleural effusion | 169 (0.48) | 0 (0.00) |
| Pulmonary hypertension | 21 (0.06) | 0 (0.00) |
| Right ventricular hypertrophy | 14 (0.04) | 0 (0.00) |
| Thromboembolic disease | 596 (1.68) | 0 (0.00) |
| Valvular disease | 625 (1.76) | 11 (0.57) |

### Appendix C Modelling

From the data, it could be seen that after the first-recorded code for breathlessness, there was a substantial number of patients who still were coded for breathlessness upon future GP consultations. This could be interpreted as prior codes being overlooked, although this is not certain. 71637 (71%) adults only had the one code for breathlessness (index date), and the remaining 29732 (29%) had breathlessness coded for again at least once during the remainder of follow-up time (median 5 years) in the cohort. The mean number of codes recorded for breathlessness across the cohort was 2 (min = 1; max = 17 (inclusive of the first-recorded code at study entry)). Out of the 29732 patients that had >1 code recorded for breathlessness in their medical record, 8834 (30%) patients did not receive a recorded diagnosis and 20898 (70%) did during follow-up. No relationship was found between repeat coding and the likelihood of receiving a recorded diagnosis subsequent to presenting with breathlessness in primary or secondary care.

**The relationship between receiving a diagnosis or not and outcomes**

136 patients died between index and landmark date and were removed from mortality models, and 24044 (23.72%) patients had a prior admission between index and landmark date. During follow-up, 47815 (47.17%) adults had an unplanned hospital admission and 11,149 (11.01%) died.

**Table C1.** Outcomes of unplanned hospital admission and all-cause mortality between adults who did and did not receive a diagnosis by landmark date.

|  | No diagnosis  (N = 57,975) | Diagnosis  (N = 43,394) |
| --- | --- | --- |
| Unplanned admission, n (%) | 24,034 (41.46) | 23,781 (54.80) |
| Death, n (%) | 4,411 (7.61) | 6,738 (15.57) |

**Causes of death**

Causes of death were investigated within 2 years of landmark date for adults presenting with breathlessness, stratified by whether a diagnosis was made or not.

**Table C2.** Causes of death within 2 years of landmark date.

| Cause of death | All | No diagnosis | Diagnosis |
| --- | --- | --- | --- |
| Number of patients, n (%) | 1,041 (100.00) | 286 (27.47) | 755 (72.53) |
| Any malignant neoplasm | 316 (30.36) | 81 (28.32) | 235 (31.13) |
| Cardiovascular disease | 302 (29.01) | 87 (30.42) | 215 (28.48) |
| Respiratory disease | 163 (15.66) | 34 (11.89) | 129 (17.09) |
| Any other cause | 139 (13.35) | 42 (14.69) | 97 (12.85) |
| Mental and behavioural disease | 52 (5.00) | 18 (6.29) | 34 (4.50) |
| Diseases of the digestive system | 42 (4.03) | 10 (3.50) | 21 (4.24) |
| Missing | 27 (2.59) | 14 (4.90) | 13 (1.72) |
